# How Price Sensitive Are Consumers? Evidence on the Demand Elasticity of Sugar-Sweetened Beverages in Bangladesh

**DOI:** 10.64898/2025.12.29.25343155

**Authors:** Abul Kalam Azad, Rumana Huque

## Abstract

The consumption of sugar-sweetened beverages (SSBs) has risen significantly in both developing and developed economies, despite well-documented evidence of their adverse health consequences. Although price-based regulations, primarily through SSB taxation, are widely recognized as a policy instrument to reduce consumption and improve public health outcomes, interventions targeting SSBs have received relatively limited attention in Bangladesh. Using the 2022 Household Income and Expenditure Survey (HIES) data and the Quadratic Almost Ideal Demand System (QUAIDS) model, we quantify own- and cross-price elasticities of SSBs to assess the responsiveness of SSB consumption to price changes. We document a substantial escalation of SSB consumption, rising from 13.2% of households in 2016 to 36.7% in 2022. The estimated uncompensated own-price elasticities are -1.060, -1.103, and -0.880 for soft drinks (liters), tea consumed outside the home (cups), and tea consumed inside the home (kilograms), respectively, suggesting pronounced price responsiveness, particularly for soft drinks and tea outside the home. Own-price elasticities for soft drinks and tea outside the home are higher among rural and low-income households compared to urban and high-income households, indicating greater sensitivity among the marginalized groups. In contrast, the elasticity estimates for tea consumed inside are higher in urban areas. The cross-price elasticity estimates suggest that soft drinks and tea consumed at home are complementary goods in the uncompensated case, while most SSB categories are found to be substitute goods across both uncompensated and compensated specifications. The findings highlight the potential effectiveness of targeted taxation policies in curbing SSB consumption, particularly soft drinks, and promoting public health in Bangladesh.

## Introduction

Intake of sugar-sweetened beverages (SSBs) poses a well-documented public health risk, contributing significantly to obesity and type 2 diabetes (Hu & Malik, 2010; Malik & Hu, 2012; Lara-Castor et al., 2025). Despite substantial evidence of their adverse health effects, consumption continues to rise at an alarming rate in both developed and developing countries. In particular, SSB consumption is rapidly expanding in developing countries, such as Bangladesh, driven by rapid urbanization and aggressive advertising by beverage companies (Azad & Huque, 2023). The Household Income and Expenditure Survey (HIES) 2016 data reveal that approximately 13.22% of households consume soft drinks, 66.77% intake sugar-added tea, coffee, and Horlicks, and 69.41% of households consume at least one sugar-added drink (Azad & Huque, 2023; Huque et al., 2024a).

SSB consumption patterns show pronounced heterogeneity across age cohorts, gender, geographic locations, occupations, and income groups. The Bangladesh Demographic and Health Survey (2022) reports that approximately 32% of children aged 6 to 23 months were given a sweetened beverage in the two days preceding data collection (NIPORT, 2023). Sugary drinks, especially soft beverages, are highly prevalent among college and university students in Bangladesh (Bipasha et al., 2017; Shahjahan et al., 2019). Singh et al. (2019) also found a high prevalence of SSB intake among this age cohort in an investigation of 187 countries. This high prevalence is primarily influenced by the low cost, pervasive marketing, attractive taste, the ubiquity of SSBs, and peer effects (Bipasha et al., 2017; Rahman et al., 2023). Additionally, studies have found that older adolescent boys consume more than younger adolescents. The former group’s consumption is observed to be higher than that of the latter group (Shamim et al., 2023).

Despite high SSB consumption, SSB regulation is less prioritized in policies and practices in Bangladesh. Empirical evidence on the effectiveness of SSB taxation and other regulatory mechanisms in curbing SSB consumption remains limited in the country. Increases in SSB prices through taxation can help curb SSB consumption. However, the magnitude of the response may vary across product types, income groups, and between rural and urban residents. Price elasticity estimates across products, income groups, and locations are necessary to design effective taxation policies for different SSBs, aiming to curb their consumption, prevent brand or product substitution, and facilitate tax modeling (Huque et al., 2024a).

The consumption of sugary drinks in Bangladesh remains relatively unexplored, with limited empirical evidence. Among the existing research, Azad and Huque (2023) investigates how SSB consumption crowds out necessary consumption, while Huque et al. (2024) examines only own-price elasticity using HIES 2016 data. On the other hand, Bipasha et al. (2017) and Rahman et al. (2023) primarily focused on the pattern of SSB consumption among a specific sample. Additionally, a steady increase in per capita income, urbanization, and demographic concentration toward the youth population have undergone significant changes, affecting how price changes influence SSB demand. We address these research gaps by employing the most recent nationally representative data available in Bangladesh and using an appropriate econometric method to generate updated, more comprehensive estimates of both own- and cross-price elasticities of SSB demand. Our examination is also disaggregated by income quantile and geographical location. The resulting elasticity estimates are expected to provide policymakers with evidence-based guidance for policy formulation and implementation, offering nuanced, distribution-sensitive insights to curb SSB consumption.

## SSB Demand Patterns and Price Elasticities in Bangladesh and Developing Economies

Bangladesh has experienced a notable increase in SSB consumption nationwide. HIES 2022 data unveils an alarming and concerning increase in the SSB consumption. We observed approximately a threefold increase in soft drink consumption from 2016 to 2022 (approximately from 13% to 37%). HIES 2022 also suggests a high and increasing volume of tea, coffee, and horlicks consumption, while 91.59% (from 69.41% in 2016) consume at least one type of sugary drink (see figure 1).

**Figure 1:**
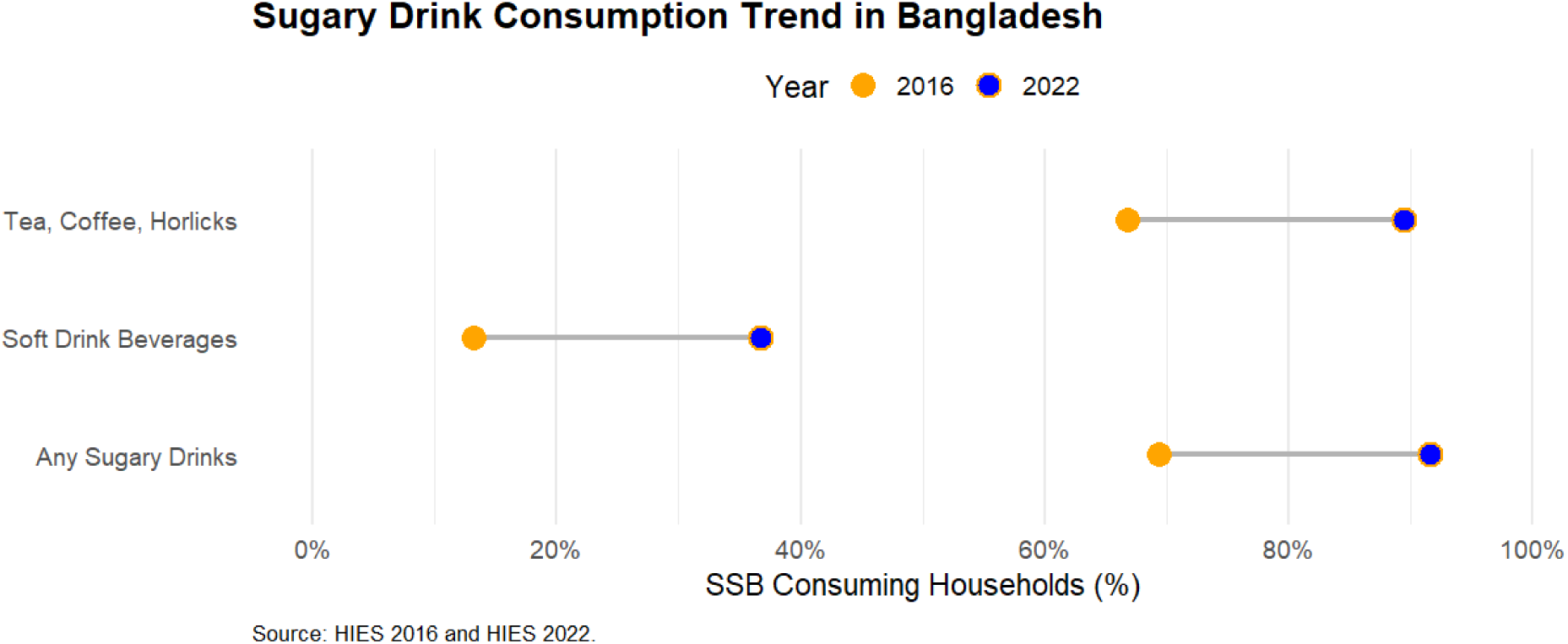
Trend of Sugary Drink Consumption in Bangladesh.

The existing global empirical literature documents cross-regional variation in price elasticity estimates for SSBs (Colchero et al., 2015; Paraje et al., 2016; Guerrero et al., 2017; Chacon et al., 2018; Maceira et al., 2018; Nor et al., 2021; John et al., 2022; Venson et al., 2023; Huque et al., 2024a). Table 1 presents the evidence of SSB price elasticities for some developing countries, including Bangladesh. Data unveils that own-price elasticity for soft drinks in Bangladesh is elastic (Huque et al., 2024a), while John et al. (2022) estimated an inelastic own-price elasticity (-0.94) for India (John et al., 2022).

**Table 1:** Price elasticities of SSBs in different countries.

|  | Years | SSBs | Soft drinks |
| --- | --- | --- | --- |
| Argentina | 2012-13 | -1.14 | - |
| Bangladesh | 2016 | - | -1.17 |
| Brazil | 2017-18 | -1.19 | - |
| Chile | 2011-12 | - | -1.37 |
| Ecuador | 2011-12 | -1.20 | - |
| Guatemala | 2014 | - | -1.39 |
| India | 2009/10 & 2011/12 | - | -0.94 |
| Malaysia | 2013-2018 | -1.11 | - |
| Mexico | 2006 | -1.10 | -1.10 |
|  | 2008 | -1.20 | -1.10 |
|  | 2010 | -1.00 | -0.90 |

Source: Colchero et al, 2015; Guerrero-López, 2017; Paraje, 2016; Venson et al., 2023; Chacon et al., 2018; John et al., 2022; Maceira et al., 2018; Nor et al., 2021; Huque et al., 2024a

In Mexico, the own-price elasticities for overall SSBs and soft drinks declined in absolute value over time. Colchero et al. (2015) reported that own-price elasticities for overall SSBs were -1.1 and -1.0 in 2006 and 2010, respectively, in Mexico (Colchero et al., 2015), while the estimates of own-price elasticity for soft drinks were -1.1 and -0.9 during the same period. Overall, the price elasticity estimates suggest a gradual decline in the absolute magnitude of own-price elasticities, indicating a shift toward increasingly inelastic consumption demand for SSBs.

## Data, Variables, and Method

### Data and variables

We employed the most recent round of Household Income and Expenditure Survey (HIES 2022) data to estimate the own and cross-price elasticities of SSBs in Bangladesh. HIES is a nationally representative dataset administered by the Bangladesh Bureau of Statistics (BBS) every five years since 1973. The BBS follows a two-stage stratified random sampling method. The primary sampling units (PSUs) were selected in the first stage, and 20 households were then randomly selected from each of these PSUs in the second stage. The detailed survey design and implementation are explained in BBS (2023).

Although no universally accepted definition for SSB exists, it commonly refers to sugary drinks and beverages. The Centers for Disease Control and Prevention (CDC) broadly defines SSBs as including soft drinks, soda, pop, soda pop, sports drinks, fruit drinks, tea, coffee, energy drinks, and sweetened milk (CDC, 2010). Zheng et al. (2015) defined tea and coffee as substitutes for SSBs, while Allcott et al. (2019), Azad & Huque (2023), and Huque et al. (2024a) encompassed sugar-added tea and coffee as the categories of SSBs. Following the CDC (2010), Allcott et al. (2019), Azad & Huque (2023), and Huque et al. (2024a), we also extended our definition of SSB by including tea consumption, as people in Bangladesh generally add sugar to their tea (Saha, 2021). However, fruit juices are mostly homemade and do not contain added sugar; hence, they are excluded from the analysis. Additionally, coffee and horlicks are also excluded due to their limited observations in the dataset.

Due to the lack of definition for SSB in HIES, we developed an operational definition of SSB and categorized them for analysis purposes. In the HIES 2022, SSBs are described under two categories: i) foods away from home, and ii) produced at home. Foods away from home include soft drinks (in milliliters) and tea (in cups), while foods at home include soft drinks (in milliliters) and tea (in kilograms). Therefore, we classified SSBs into three categories: i) total soft drink consumption by aggregating their home and away-from-home consumption (in liters), ii) tea consumed away from home (in cups), and iii) tea consumed at home (in kilograms).

### Empirical model

We model consumer behaviour using a utility-based model to determine the price effects on consumer demand. In particular, we applied the Quadratic Almost Ideal Demand System (QUAIDS) model (Banks et al., 1997), which permits greater flexibility than the traditional income-expenditure (Engel) curves. Demand system analysis models households’ expenditure behaviour across a set of related goods to estimate own-and cross-price elasticities. However, empirical investigation of such a demand system often encounters methodological challenges. For instance, the law of demand posits an inverse relationship between quantity demanded and price, though they are jointly determined in most cases in the real world. This joint determination often results in identification and endogeneity issues. The failure to address these econometric issues may often lead to biased and inconsistent parameter estimation (John et al., 2023).

To address the identification or endogeneity problem, the Almost Ideal Demand System (AIDS) and its quadratic extension, Quadratic AIDS (QUAIDS) model, are widely used among researchers to estimate own and cross-price elasticities (Deaton & Muellbauer, 1980). However, we adopt the QUAIDS model instead of the AIDS model, as the QUAIDS model has greater flexibility than the AIDS model. Additionally, the AIDS model is nested within the QUAIDS model. Banks et al. (1997) used the following indirect utility function to estimate the QUAIDS model. By following Banks et al. (1997) and Poi (2012), we model as follows:

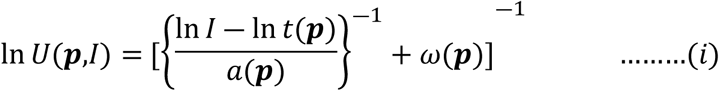

where, 𝑈(𝑝,𝐼) is the indirect utility function, 𝒑 is the price vector, 𝐼 is income or expenditure, and ln 𝑡(𝒑) is the transcendental logarithm function in equation (ii). Moreover, 𝑎(𝒑) in equation (iii) is the price aggregator in Cobb-Douglas form, whereas 𝜔(𝒑) is described in equation (iv):

𝑙𝑛 𝑡(𝒑) = 𝜑

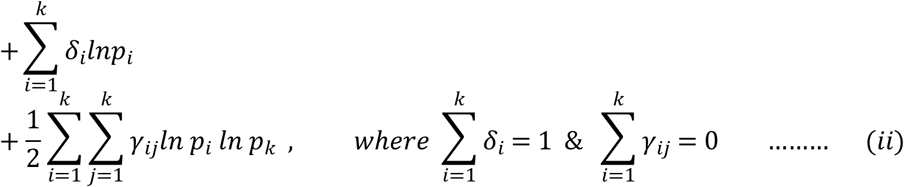

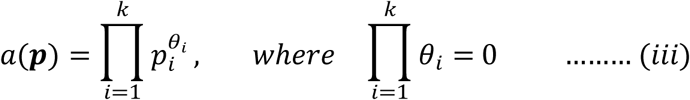

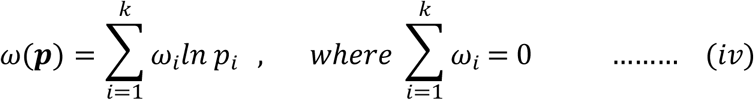

for all 𝑖, 𝑗 = 1, ………, 𝑘. Banks et al. (1997) define equations (i), (ii), (iii), and (iv) combinedly as the QUAIDS system. Although the parameters 𝛿_𝑖_, 𝛾_𝑖𝑗_, 𝜃_𝑖_, and 𝜂_𝑖_parameters need to be estimated, however, the QUAIDS demand system requires 𝜑 parameter to be assumed a value by the discretion of the researcher. Estimating the QUAIDS demand system also requires Slutsky’s symmetry and homogeneity in its estimation process.

Assume 𝑐_𝑖_ is the expenditure share or budget share of good 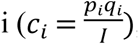, where 𝑞*_i_* is the amount of quantity demanded of good 𝑖 by a household and 𝐼 is the total expenditure on the particular group. Hence, the expenditure share of good 𝑖 can be derived from the equation 𝑖 using Roy’s identity as follows:

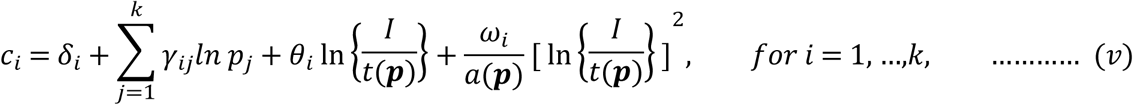

Equation (v) clearly drops the quadratic expression from the expenditure share equation and turns into the following linear equation when 𝜔_𝑖_ = 0:

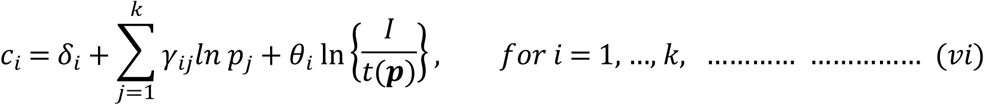

This linear equation is Deaton and Muellbauer’s (1980) original Almost Ideal Demand System (AIDS) equation. Although equation (vi) requires nonlinear estimation due to the price indices, Deaton and Muellbauer’s (1980) assumed a linear approximation for 𝑙𝑛 𝑡(𝑝). We can also test the null hypothesis 𝐻_0_: 𝜔_𝑖_ = 0 to determine whether we should use the AIDS or the QUAIDS model. The statistically significant value of 𝜔_𝑖_allows us to choose the QUAIDS demand system model over the AIDS model.

The QUAIDS demand system may also encompass household-specific characteristics developed by Ray (1983) and strengthened by Poi (2002). Following Poi (2012), if the expenditure function is assumed to take the form e(𝐩,𝐱,u) = 𝑚_0_(𝒑, 𝒙,𝑢) ∗ 𝑒(𝒑,𝑢), Ray (1983) decomposes the scaling function in the following technique 𝑚_0_(𝒑, 𝒙,𝑢) = 𝑚_0_(𝒙) ∗ 𝜇(𝒑, 𝑢, 𝒙) and 𝑚_0_(𝒙) = 1 + 𝜌′𝒙 . Where **x** is the vector of household-specific characteristics (household size, number of adult members, household head education, gender, age, having a refrigerator, remittance-receiving status, access to the internet, and geographical location). After including the household characteristics, we can present the household expenditure share equation as follows:

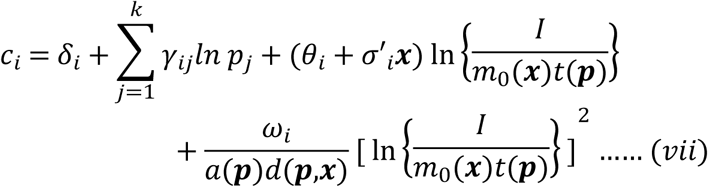

where, 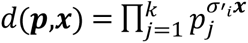

By following Poi (2012), we can model the Marshallian (uncompensated) price elasticity and expenditure or income elasticity for good 𝑖, respectively, as follows:

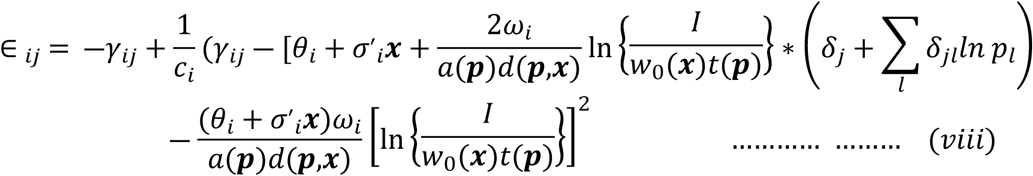

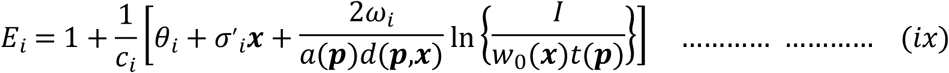

Equations (viii) and (ix) are attained through the derivation of equation (vii) with respect to the unit value or price and expenditure, respectively. Equations (viii) and (ix) offer the price and expenditure elasticities, respectively. We can also derive the Hicksian (compensated) price elasticities (both own- and cross-price elasticities) using the Slutsky equation: 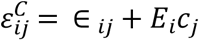 (Poi, 2012).

## Results

To characterize the households by SSB consumption patterns, we classified them into four groups based on their intake of sugary drinks: (i) households with any SSB consumption, (ii) households consuming soft drinks, (iii) households consuming tea outside the home, and (iv) households with tea inside the home (Table 2). We found that any form of sugary drink consumption is associated with substantially higher income and expenditure compared with households with no sugary drink consumption (Table 2). Particularly, households that consume soft drinks monthly earn BDT 42, 212 (=$460.33)^1^ and spend BDT 39,687 ($432.79) on average across the categories, while these estimates of monthly income and expenditure are approximately BDT 19,134 (=$208.66) and BDT 17,863 (=$194.80), respectively, for households without SSB consumption. On the other hand, the income and expenditure for households consuming tea inside and outside the home are quite similar. This finding aligns with the findings obtained by Azad and Huque (2023).

**Table 2:** Descriptive statistics based on household drinking status.

|  | Household Categorization based on their drinking status |  |  |  |  |
| --- | --- | --- | --- | --- | --- |
| Variables | Households without any sugary drinks | Households with any sugary drinks | Households with soft drinks | Households with tea outside | Households with tea inside |

<sup>1</sup> Considering 1 USD = 91.7 BDT; Source: World Bank [Official exchange rate BDT/USD downloaded 12/27/2025]

|  |  |  |  |  |  |
| --- | --- | --- | --- | --- | --- |
| Household Monthly Income (BDT) | 19134.740<br>(15469.990, 22799.500) | 33377.580<br>(32119.080, 34636.070) | 42212.140<br>(39959.700, 44464.590) | 31933.640<br>(30577.700, 33289.580) | 32203.160<br>(31007.930, 33398.380) |
| Household Monthly Expenditure (BDT) | 17862.760<br>(17068.160, 18657.360) | 31435.190<br>(31008.470, 31861.900) | 39687.320<br>(38867.820, 40506.820) | 30528.240<br>(30101.440, 30955.040) | 30316.050<br>(29914.390, 30717.700) |
| Household size | 3.623<br>(3.526, 3.721) | 4.398<br>(4.368, 4.428) | 4.529<br>(4.480, 4.577) | 4.425<br>(4.392, 4.458) | 4.334<br>(4.306, 4.363) |
| Number of adult members | 2.328<br>(2.267, 2.389) | 2.898<br>(2.876, 2.919) | 2.945<br>(2.909, 2.981) | 2.931<br>(2.908, 2.955) | 2.851<br>(2.830, 2.871) |
| Education level of household head | 2.353<br>(2.111, 2.594) | 2.946<br>(2.843, 3.048) | 3.201<br>(3.035, 3.368) | 2.750<br>(2.652, 2.849) | 2.897<br>(2.801, 2.993) |
| Gender (=1 if household head is male, otherwise 0) | 0.704<br>(0.678, 0.730) | 0.900<br>(0.895, 0.905) | 0.882<br>(0.873, 0.891) | 0.931<br>(0.926, 0.935) | 0.884<br>(0.878, 0.889) |
| Age in years | 47.392<br>(46.529, 8.255) | 46.920<br>(46.689, 47.150) | 45.775<br>(45.422, 46.128) | 46.895<br>(46.644, 47.147) | 46.959<br>(46.736, 47.182) |
| Refrigerator (=1 if a household has, otherwise 0) | 0.355<br>(0.328, 0.383) | 0.553<br>(0.545, 0.562) | 0.661<br>(0.649, 0.674) | 0.529<br>(0.519, 0.538) | 0.537<br>(0.529, 0.545) |
| Remittance (=1 if received, otherwise 0) | 0.168<br>(0.146, 0.189) | 0.147<br>(0.141, 0.153) | 0.173<br>(0.163, 0.183) | 0.122<br>(0.115, 0.128) | 0.149<br>(0.143, 0.155) |
| Internet access (=1 if a household has internet access, otherwise 0) | 0.457<br>(0.429, 0.486) | 0.686<br>(0.678, 0.694) | 0.781<br>(0.769, 0.792) | 0.672<br>(0.663, 0.680) | 0.667<br>(0.659, 0.675) |
| Location | 0.363<br>(0.336, 0.39062) | 0.512<br>(0.503, 0.520) | 0.567<br>(0.553, 0.580) | 0.501<br>(0.492, 0.511) | 0.500<br>(0.491, 0.508) |
| Observations | 1,176 | 13086 | 5256 | 10784 | 14262 |

Note: 95% confidence interval is in the parentheses

Household heads in SSB-consuming households exhibit the largest level of educational attainment. The number of adult members is also higher for households that consume SSBs. However, the number is broadly comparable across the SSB-consuming groups, ranging from 2.85 to 2.95 on average. Sugary drinks are more popular among households headed by younger individuals and males. This SSB consumption pattern suggests that older and female individuals in the country are less inclined towards SSB consumption. Moreover, households living in urban areas with greater asset ownership, such as refrigerators, and higher internet access, consume more SSBs than households without SSB intake (Table 2).

All elasticity coefficients are found statistically significant at a 1% level. The findings indicate that a 10% increase in the price of soft drinks results in an average decrease of approximately 10.6% in consumption demand, holding other factors constant. The result implies that soft drinks exhibit price-elastic demand in Bangladesh.

**Table 3:** Own Price Elasticity of SSBs.

| SSBs | Uncompensated (Marshallian) price elasticities |  | Compensated (Hicksian) price elasticities |  |
| --- | --- | --- | --- | --- |
|  | Price elasticities | 95% conf. interval | Price elasticities | 95% conf. interval |
| Soft drinks (Liter) | -1.060***<br>(0.042) | [-1.142, -0.977] | -0.660***<br>(0.043) | [-0.744, -0.577] |
| Tea Outside (Cup) | -1.103***<br>(0.038) | [-1.178, -1.028] | -0.687***<br>(0.037) | [-0.759, -0.615] |
| Tea Inside (KG) | -0.880***<br>(0.057) | [-0.992, -0.768] | -0.695***<br>(0.055) | [-0.804, -0.586] |

Notes: \*\*\*p < 0.01; Standard errors are in parentheses

For tea consumed outside the home, a 10% price increase on a tea cup reduces the demand for tea outside the home by 11.03% on average. The elastic price elasticity of tea consumption that, holding other factors constant, tea consumption outside the home is highly responsive to price changes. In contrast, the uncompensated own-price elasticity of inside-home tea consumption is inelastic, with a magnitude of -0.880. This finding suggests that tea consumption within the home is likely a necessity. The finding aligns with the expectation, as households are more likely to adjust their outside home consumption before reducing the inside home consumption of the same goods.

As expected, the compensated (Hicksian) price elasticities are smaller in absolute value than the uncompensated (Marshallian) elasticities. The compensated own-price elasticities for soft drinks, tea consumed outside, and tea consumed inside the home are -0.660, -0.687, and -0.695, respectively. Consistent with the uncompensated estimates, the compensated elasticities are also highly statistically significant. The attenuation in magnitude of compensated price elasticities reflects theoretical differences. The compensated price elasticities only capture the substitution effect of the price change, holding utility constant, while the uncompensated price elasticities encompass both the substitution and income effects, providing a comprehensive categorization of price changes in consumer demand.

The estimate of cross-price elasticities of SSBs indicates that an increase in the price of tea cups consumed outside leads to an increase in SSB consumption, suggesting they are substitute goods. The magnitude of the cross-price elasticity is 0.082, implying that a 10% increase in the price of tea per cup outside the home increases the consumption demand for soft drinks by about 8.2%. This result is particularly noteworthy because a high price of tea outside the home may induce consumers to shift into soft drinks. Similarly, an increase in the price of soft drinks leads to an increase in the demand for tea consumption outside, although the coefficient is not statistically significant, implying a weaker substitution in this direction. Additionally, tea consumption at home and away from home appears as a substitute good - a 10% increase in the price of a tea cup leads to a 0.5% increase in inside tea demand, and a 10% increase in the price of inside tea increases the demand for outside tea by 1.84% (see Table 4). On the other hand, the consumption of soft drinks and tea inside the home is found to be complementary goods. A 10 % increase in the price of inside tea consumption causes a decrease in soft drinks consumption by about 1.0%, and a 10% increase in the price of soft drinks decreases inside tea consumption by about 1.8%.

**Table 4:** Cross-Price Elasticity of SSBs.

| SSBs | Uncompensated cross-price elasticities |  |  | Compensated price elasticities |  |  |
| --- | --- | --- | --- | --- | --- | --- |
|  | Soft drinks (Liter) | Tea Outside (Cup) | Tea Inside (KG) | Soft drinks (Liter) | Tea Outside (Cup) | Tea Inside (KG) |
| Price of Soft drinks (Liter) | - | 0.009<br>(0.041) | -0.180***<br>(0.032) | - | 0.543***<br>(0.037) | 0.118***<br>(0.030) |
| Price of Tea Outside (Cup) | 0.082***<br>(0.029) | - | 0.050*<br>(0.027) | 0.402***<br>(0.028) | - | 0.286***<br>(0.026) |
| Price of Tea Inside (KG) | -0.099**<br>(0.042) | 0.184***<br>(0.050) | - | 0.170***<br>(0.042) | 0.525***<br>(0.049) | - |

Notes: \* $p < 0.10$ ; \*\* $p < 0.05$ ; \*\*\* $p < 0.01$ ; Standard errors are in parentheses

Regarding the compensated cross-price elasticities, some variations emerge in the signs and significance level of the cross-price elasticity coefficients. Although the uncompensated cross-price elasticities indicate that soft drinks and inside tea consumption are complementary goods, the compensated cross-price elasticities present these two sugary drinks as substitute goods. In addition, all the cross-price elasticity coefficients are highly statistically significant, including the elasticity coefficient of outside tea consumption with respect to soft drinks, which was not statistically significant in the uncompensated estimation (see Table 4). Despite the deviations between uncompensated and compensated cross-price elasticities, the general pattern remains consistent across both specifications, suggesting that sugary drinks are mostly substitutes.

In rural areas, the obtained elasticities are 1.081, -1.114, and -0.847 for soft drinks, tea consumed outside the home, and tea consumed inside the home, respectively (see Table 5). These estimates suggest that a 10% price increase is associated with a 10.81%, 11.14%, and 8.47% decrease in their consumption, respectively. In urban areas, the corresponding uncompensated price elasticities are -1.047, -1.097, and -0.899 for soft drinks, tea consumed, and inside the home, respectively. For soft drinks and tea consumed outside the home, the absolute estimates of elasticities are smaller in urban areas than in rural regions, whereas the elasticity estimate for tea consumed at home is marginally larger in urban areas. Overall, the greater absolute magnitudes of price elasticity in the rural areas hint that rural consumers exhibit greater sensitivity to price changes in consumption demand for soft drinks and tea consumed outside. In contrast, urban households appear to be more price-sensitive in their demand for in-home tea consumption.

**Table 5:** Uncompensated and Compensated for Own Price Elasticity of SSBs for Rural and Urban Regions.

| SSBs | Uncompensated price elasticities |  | Compensated price elasticities |  |
| --- | --- | --- | --- | --- |
|  | Rural | Urban | Rural | Urban |
| Soft drinks (Liter) | -1.081***<br>(0.049) | -1.047***<br>(0.042) | -0.689***<br>(0.049) | -0.644***<br>(0.042) |
| Tea Outside (Cup) | -1.114<br>(0.040)*** | -1.097***<br>(0.039) | -.686***<br>(0.038) | -0.688***<br>(0.038) |
| Tea Inside (KG) | -0.847***<br>(0.057) | -0.899***<br>(0.058) | -0.668***<br>(0.055) | -0.711***<br>(0.057) |

Notes: \*\*\*p < 0.01; Standard errors are in parenthesis

On the other hand, the corresponding compensated own-price elasticity estimates for soft drinks, tea consumed outside, and inside the home for rural (urban) areas are -0.689, -0.686, and -0.668 (-0.644, -0.688, and -0.711), respectively. Consistent with the overall elasticity estimates (Table 3), the estimated compensated own price elasticities are also smaller than the uncompensated elasticities for rural areas, highlighting the predominance of the substitution effect.

Tables A1a and A1b in the Appendix present the uncompensated and compensated cross-price elasticities for rural and urban regions, respectively. The signs and significance levels are broadly consistent in both cases. For both uncompensated and compensated elasticities, soft drinks and tea consumed outside the home are identified as substitute goods in both rural and urban areas. On the other hand, soft drinks and tea consumed at home are found to be complementary goods under uncompensated price elasticities, whereas they are estimated as substitute goods under compensated price elasticities for both rural and urban areas. Additionally, tea, whether consumed at home or outside, consistently emerges as a substitute for all specifications.

The own price elasticities across the income quantiles suggest that all coefficients of price elasticities are statistically significant at the 1% level. As household income increases, the absolute magnitudes of uncompensated own-price elasticities for soft drinks and tea inside the home decrease sharply, making sugary drinks increasingly less elastic goods for the higher-income group. For instance, a 10% increase in the price of soft drinks leads to a decrease of about 11.63% on average for the lowest income quantile, while a 10.34% decrease is observed for the top income quantile group. Regarding the elasticity of tea consumption outside the home, these elasticities range from -1.122 to -1.076, from the first quantile to the fifth quantile. The findings reveal greater price responsiveness among the lower-income quantile groups for soft drinks and tea outside the home. Additionally, all uncompensated price elasticities for both soft drinks and outside tea consumption exhibit elastic demand across all income groups. However, we found a mostly reverse pattern of uncompensated own price elasticities for tea consumed at home, ranging from -0.848 to -0.913 from the first to the fifth income quantile. In addition, we found slight variations in elasticity estimates among the middle three income quantiles for all sugary drinks, meaning that the middle-income groups exhibit substantially consistent behavior regarding SSB consumption.

**Table 6:** Uncompensated and Compensated Own Price Elasticity of SSBs based on Income Quantiles.

|  | Uncompensated price elasticities |  |  | Compensated price elasticities |  |  |
| --- | --- | --- | --- | --- | --- | --- |
|  | Soft drinks | Tea Outside | Tea Inside | Soft drinks | Tea Outside | Tea Inside |
| **1st Income Quantile** | -1.163***<br>(0.088) | -1.122***<br>(0.044) | -0.848***<br>(0.062) | -0.781***<br>(0.087) | -0.677***<br>(0.040) | -0.675***<br>(0.059) |
| **2nd Income Quantile** | -1.054***<br>(0.043) | -1.130***<br>(0.042) | -0.873***<br>(0.060) | -0.671***<br>(0.044) | -0.688***<br>(0.038) | -0.697***<br>(0.058) |
| **3rd Income Quantile** | -1.068***<br>(0.043) | -1.120***<br>(0.039) | -0.858***<br>(0.057) | -0.680<br>(0.043) | -0.686***<br>(0.037) | -0.680***<br>(0.055) |
| **4th Income Quantile** | -1.049***<br>(0.042) | -1.103***<br>(0.038) | -0.870***<br>(0.057) | -0.653***<br>(0.043) | -0.682***<br>(0.036) | -0.687***<br>(0.055) |
| **5th Income Quantile** | -1.034***<br>(0.043) | -1.076***<br>(0.043) | -0.913***<br>(0.060) | -0.613***<br>(0.044) | -0.695***<br>(0.041) | -0.715***<br>(0.059) |

Notes: \*\*\*p < 0.01; Standard errors are in parenthesis

Consistent with uncompensated elasticities, the compensated own price elasticities depict similar patterns for SSB consumption across income quantiles. For the soft drinks, the compensated own-price elasticity for the first quantile is -0.781, and for the fifth quantile is -0.613, indicating that a 10% price increase reduces soft drink consumption by 7.81% and 6.13% for the first and fifth quantiles, respectively. This gradient exhibits greater responsiveness among the lower-income groups. Regarding the outside and inside tea consumption, we find an increasing pattern of elasticity magnitudes in absolute value. For instance, a 10% price increase causes a 6.77% reduction for the first income quantile, while a 6.95% reduction for the fifth income quantile. On the other hand, the compensated price elasticities for inside tea consumption are -0.675, -0.697, - 0.680, -0.687, and -0.715, respectively, from the first to the fifth quantiles. As with uncompensated elasticities, we also find little variation among the middle three income quantiles for all sugary drinks, suggesting a broadly homogeneous consumption behavior for SSBs among middle-income groups.

Tables A2a-A2e in the Appendix report the uncompensated and compensated cross-price elasticities from the first to the fifth income quantiles, respectively. Overall, the signs and magnitudes across income groups are broadly consistent with only minor deviations from the overall cross-price elasticities depicted in Table 4. Across household income groups, soft drinks and tea consumed outside the home generally exhibit a substitute relationship. However, an exception appears in the fifth income quantile (Table A2e), where the uncompensated cross-price elasticity for soft drinks and tea consumed outside is negative, though the estimate is not statistically significant.

All estimates of expenditure elasticities for sugary drinks are found positive and statistically significant at the 1% level (see Table 7), implying that consumption demand for these beverages rises with household expenditure. The estimated expenditure elasticities indicate that a 10% increase in household expenditure leads to an increase in consumption of 11.95%, 9.90%, and 7.53 for soft drinks, tea consumed inside and outside the home, respectively. The elastic magnitude of the soft drink underscores its classification as a luxury good.

**Table 7:** Expenditure Elasticity of SSBs.

| SSBs | Expenditure elasticities<br>(Coefficients) | [95% conf. interval] |
| --- | --- | --- |
| Soft drinks (Liter) | 1.195***<br>(0.035) | [1.126, 1.263] |
| Tea Outside (Cup) | 0.990***<br>(0.027) | [0.937, 1.043] |
| Tea Inside (KG) | 0.753***<br>(0.040) | [0.675, 0.831] |

Notes: \*\*\* $p < 0.01$ ; Standard errors are in parentheses. Elasticities are calculated at prices, demographic variables, and expenditure means.

We carry out a likelihood-ratio (LR) test under the null hypothesis that the quadratic term of the QUAIDS specification is zero (𝐻_0_: 𝜔_𝑖_ = 0), which corresponds to the AIDS model being nested within QUAIDS to assess the appropriate functional form of the demand system. The obtained LR estimate is 4.90 and statistically significant, meaning that the statistically significant likelihood-ratio test suggests that the QUAIDS specification better fits the data compared to the AIDS specification. We also plot the budget share of each SSB category against total expenditure to explore the relationship between household spending and SSB consumption. As portrayed in panels (a)-(c) of Figure 2, a nonlinear relationship between (log of) total expenditure and SSB categories is observed. This visual evidence is largely consistent with our quadratic hypothesis (QUAIDS model).

**Figure 2:**
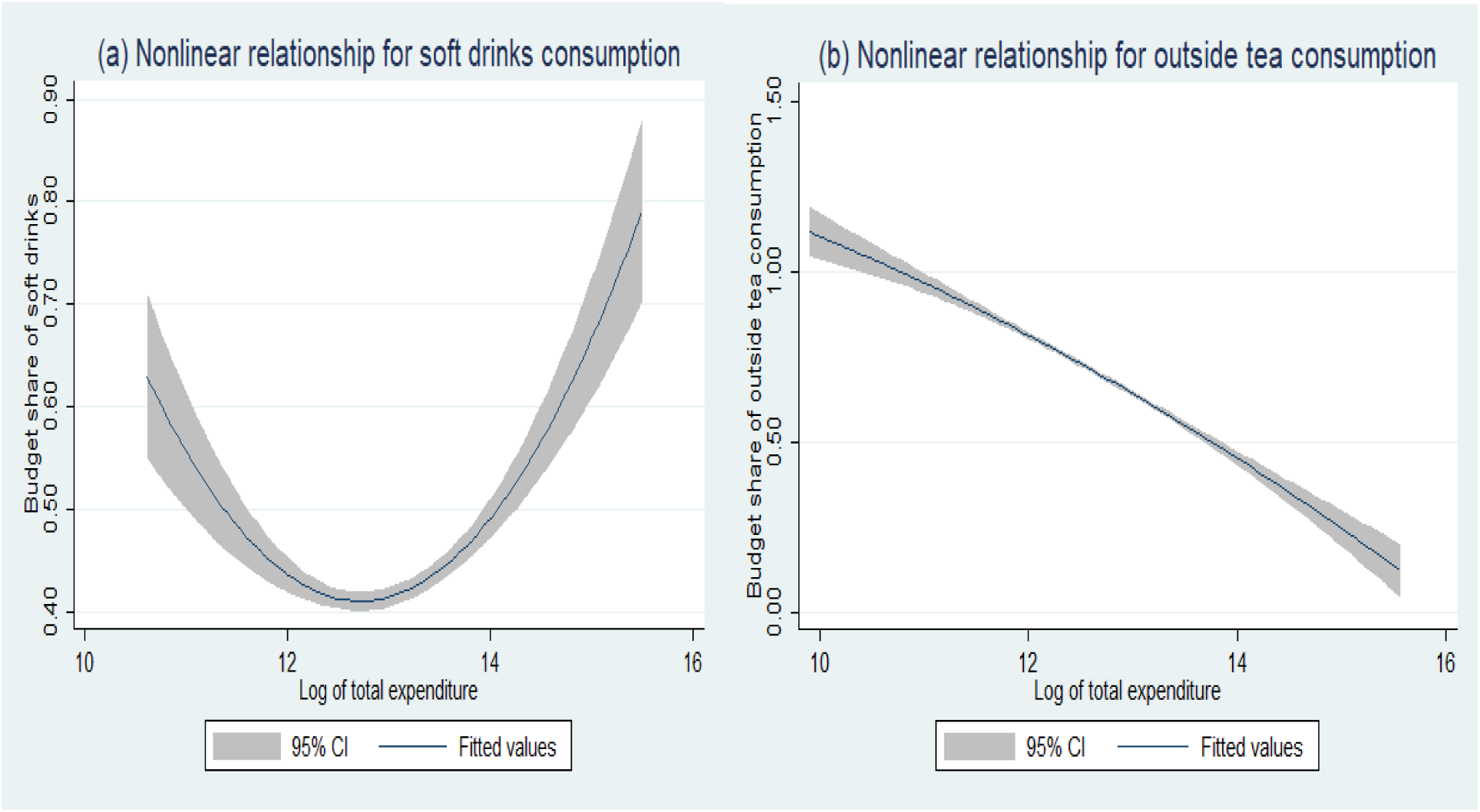
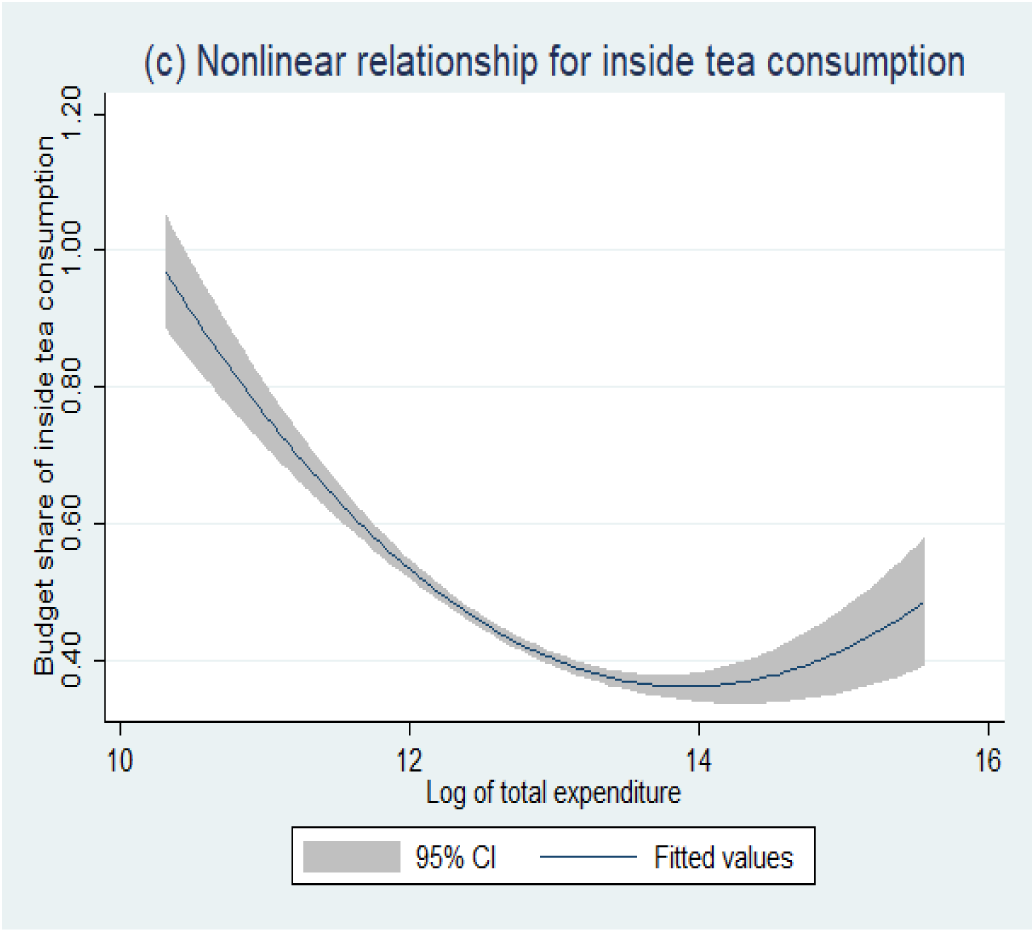
Nonlinear relationship between budget share and (ln) expenditure. Source: Authors generated the graph from the data

## Discussion

Our findings suggest that the proportion of households consuming soft drinks increased about threefold between 2016 (13.22%) and 2022 (36.70%) in Bangladesh, posing a serious public health threat. Additionally, our estimates indicate that the consumption of other sugary drinks has increased substantially over the last decade. These trends underscore the urgency of a considerable increase in SSB prices through efficient taxation to curb SSB consumption.

Our results reveal that the uncompensated own-price elasticity estimates for the demand for soft drinks, tea consumed outside, and inside the home are -1.060, -1.103, and -0.880, respectively. We also find that the demand for soft drinks and tea consumed outside is more elastic among households living in rural areas, those in more marginalized municipalities, and those with lower incomes. Huque et al. (2024a) found that the own-price elasticity of soft drinks is about -1.17 in 2016, indicating that the responsiveness of price changes has decreased over time. Additionally, soft drink consumption has not only increased three times, but has also become more inelastic over time. These findings of the price elasticity coefficient for soft drinks may result from increased habituation or market penetration. This development might have a profound negative impact on public health.

The estimated elastic price elasticity coefficients suggest that increasing taxes on SSBs can considerably reduce SSB consumption while simultaneously increasing government revenue. A large and growing number of studies document the health benefit measured by the number of lives saved through taxation on SSBs (Ward & Gortmaker, 2020; Krieger et al., 2021; Popkin & Ng, 2021). Thus, the findings of this study reinforce the importance of the progressive nature of SSB taxation for achieving the dual policy goals of improving public health and increasing government tax revenue.

Comparing the estimations across income groups, it is evident that individuals in the low-income group exhibit, on average, greater responsiveness to price changes than their high-income counterparts. Thus, an inverse relationship between SSB price and its demand across socioeconomic and demographic groups indicates that lower- and middle-income households would reduce SSB consumption more substantially in response to price changes. In other words, the greater absolute magnitude of the elasticity parameter for the lower-income and middle-income groups implies that, in response to price increases, members of those income groups will reduce their consumption more than the top-income group.

The effectiveness of SSB taxation is largely dependent on the efficiency of the tax structure design and the proper implementation of tax policy. However, SSB taxation is less researched in Bangladesh. The tax structure should be designed to limit the incentive to switch to lower-priced products. Moreover, the tax must be annually adjusted for inflation and income growth to maintain its effectiveness in relation to the real value of the SSB price. Non-price measures, such as regulating aggressive industry marketing and increasing mass awareness of the population about the harmful effects of SSB consumption, are also important for reducing SSB consumption. The tax administration also needs to be strengthened to achieve the dual purposes of protecting public health goals and increasing the government’s revenue stream (Huque et al., 2024b).

The study has some limitations. Since there is no precise definition of SSBs among the practitioners in the SSB field, we define SSBs for operational purposes. We only encompassed all sugar-added soft drinks and tea consumed inside and outside the home, but we excluded homemade fruit juice, which we assume is mostly sugar-free. Additionally, we excluded coffee-consuming households from the study due to the limited observations available. Third, although some consumers are accustomed to consuming sugar-free tea, we were unable to segregate these households from the sample due to the nature of the HIES data.

## Conclusion

The consumption of SSBs in Bangladesh has increased steadily over time, placing an additional burden on the existing, constrained public health budget by diverting scarce resources away from primary healthcare services. The increasing challenges and burden on public health underscore the importance of curbing SSB consumption through pragmatic and evidence-based policy formulation. Identifying price elasticities is immensely useful for evidence-based policy purposes, especially for the taxation on the price of sugary drinks.

The existing literature on SSBs in Bangladesh is limited. We found only one study (Huque et al., 2024a) that focused on identifying price elasticities. However, that study was only based on past HIES 2016 data and focused exclusively on own-price elasticities in Bangladesh. This study contributes to the literature by offering the first comprehensive estimate of own and cross-price elasticities, taking into account regional and income variations, utilizing the latest available data. We observe that the price elasticity for soft drinks is relatively smaller in absolute value compared to the estimate of Huque et al. (2024), implying that soft drinks have been increasingly perceived as quasi-necessity goods. Therefore, it calls for a cautious and well-calibrated approach to taxation policy on the prices of sugary drinks, aiming to mitigate the public health burden associated with the rising consumption of sugary drinks.

## Declarations

### Ethics approval and consent to participate

We have analyzed secondary data from a nationally representative survey. Hence, ethical clearance was not required to carry out the analysis.

### Consent for publication

Not applicable.

### Availability of data and materials

The datasets used and/or analyzed during the current study are available from the Household Income and Expenditure Survey, Bangladesh - [; web link: http://www.bbs.gov.bd/] on reasonable request.

## Acknowledgment

Not applicable

## Appendix

**Table A1a:** Uncompensated and Compensated Cross Price Elasticity of SSBs for Rural Region.

| SSBs | Uncompensated Cross Price Elasticity |  |  | Compensated Cross Price Elasticity |  |  |
| --- | --- | --- | --- | --- | --- | --- |
|  | Soft drinks | Tea Outside | Tea Inside | Soft drinks | Tea Outside | Tea Inside |
| Soft drinks (Liter) | - | 0.023<br>(0.045) | -0.206***<br>(0.034) | - | 0.575 ***<br>(0.042) | 0.113***<br>(0.033) |
| Tea Outside (Cup) | .083***<br>(0.029) | - | (0.028<br>(0.027) | 0.405***<br>(0.029) | - | 0.281***<br>(0.026) |
| Tea Inside (KG) | -0.094**<br>(0.041) | 0.182***<br>(0.050) | - | 0.157***<br>(0.041) | 0.511***<br>(0.048) | - |

Notes: \*\*\*p < 0.01; Standard errors are in parenthesis

**Table A1b:**
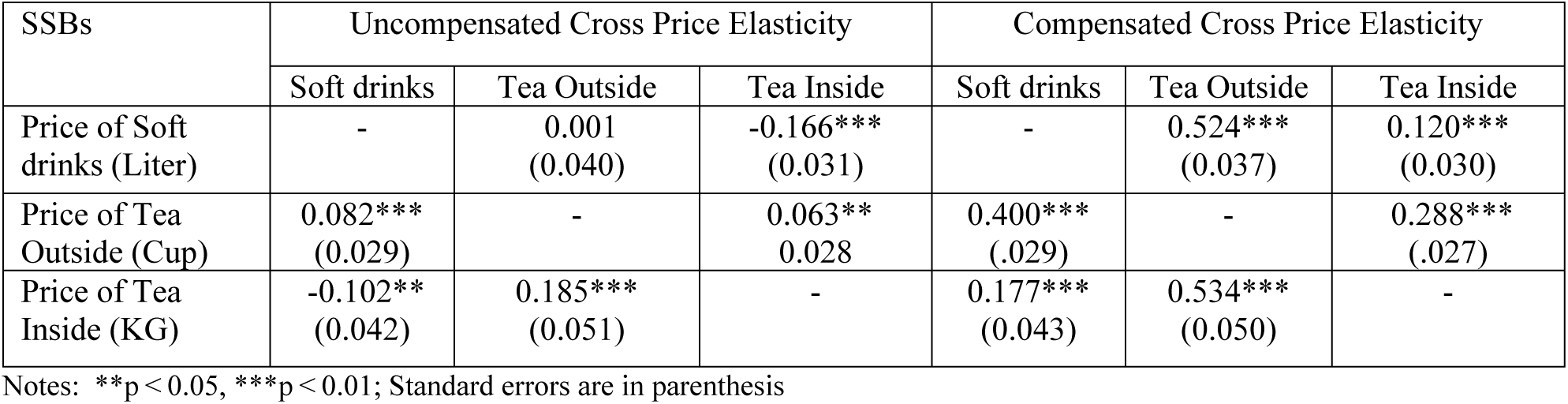
Uncompensated and Compensated Cross-Price Elasticity of SSBs for Urban Region.

**Table A2a:** Uncompensated and Compensated Cross-Price Elasticity of SSBs for 1^st^ Income Quantile.

| SSBs | Uncompensated Cross Price Elasticity |  |  | Compensated Cross Price Elasticity |  |  |
| --- | --- | --- | --- | --- | --- | --- |
|  | Soft drinks | Tea Outside | Tea Inside | Soft drinks | Tea Outside | Tea Inside |
| Price of Soft drinks (Liter) | - | 0.044<br>(0.058) | -0.239***<br>(0.042) | - | 0.673***<br>(0.067) | 0.108**<br>(0.043) |
| Price of Tea Outside (Cup) | 0.084***<br>(0.030) | - | 0.026 (0.028) | 0.404***<br>(0.030) | - | 0.273***<br>(0.027) |
| Price of Tea Inside (KG) | -0.094**<br>(0.042) | 0.191***<br>(0.056) | - | 0.151***<br>(0.043) | 0.0524***<br>(0.052) | - |

Notes: \*\*p < 0.05, \*\*\*p < 0.01; Standard errors are in parentheses

**Table A2b:** Uncompensated and Compensated Cross-Price Elasticity of SSBs for 2^nd^ Income Quantile.

| SSBs | Uncompensated Cross Price Elasticity |  |  | Compensated Cross Price Elasticity |  |  |
| --- | --- | --- | --- | --- | --- | --- |
|  | Soft drinks | Tea Outside | Tea Inside | Soft drinks | Tea Outside | Tea Inside |
| Price of Soft drinks (Liter) | - | 0.036<br>(0.045) | -0.171***<br>(0.033) | - | 0.562***<br>(0.039) | 0.109***<br>(0.031) |
| Price of Tea Outside (Cup) | 0.082***<br>(0.029) | - | 0.039<br>(0.027) | 0.408***<br>(0.029) | -- | 0.280***<br>(0.026) |
| Price of Tea Inside (KG) | -0.097**<br>(0.043) | 0.199***<br>(0.055) | - | 0.156***<br>(0.044) | 0.541***<br>(0.052) | - |

Notes: \*\*p < 0.05, \*\*\*p < 0.01; Standard errors are in parentheses

**Table A2c:** Uncompensated & Compensated Cross-Price Elasticity of SSBs for 3^rd^ Income Quantile.

| SSBs | Uncompensated Cross Price Elasticity |  |  | Compensated Cross Price Elasticity |  |  |
| --- | --- | --- | --- | --- | --- | --- |
|  | Soft drinks | Tea Outside | Tea Inside | Soft drinks | Tea Outside | Tea Inside |
| Price of Soft drinks (Liter) | - | 0.029<br>(0.044) | -0.188***<br>(0.033) | - | 0.568***<br>(0.039) | 0.112***<br>(0.031) |
| Price of Tea Outside (Cup) | 0.082***<br>(0.028) | - | 0.035<br>(0.027) | 0.405***<br>(0.028) | - | 0.281***<br>(0.026) |
| Price of Tea Inside (KG) | -0.096**<br>(0.041) | 0.189***<br>(0.051) | - | 0.157***<br>(0.042) | 0.523***<br>(0.049) | - |

Notes: \*\*p < 0.05, \*\*\*p < 0.01; Standard errors are in parentheses

**Table A2d:** Uncompensated and Compensated Cross-Price Elasticity of SSBs for 4^th^ Income Quantile.

| SSBs | Uncompensated Cross Price Elasticity |  |  | Compensated Cross Price Elasticity |  |  |
| --- | --- | --- | --- | --- | --- | --- |
|  | Soft drinks | Tea Outside | Tea Inside | Soft drinks | Tea Outside | Tea Inside |
| Price of Soft drinks (Liter) | - | 0.007<br>(0.041) | -0.179***<br>(0.032) | - | 0.538***<br>(0.037) | 0.115***<br>(0.030) |
| Price of Tea Outside (Cup) | 0.082***<br>(0.028) | - | 0.045*<br>(0.027) | 0.400***<br>(0.028) | - | 0.282***<br>(0.026) |
| Price of Tea Inside (KG) | -0.098**<br>(0.041) | 0.182***<br>(0.050) | - | 0.163***<br>(0.042) | 0.524***<br>(0.048) | - |

Notes: \*p < 0.10, \*\*p < 0.05, \*\*\*p < 0.01; Standard errors are in parentheses

**Table A2e:** Uncompensated and Compensated Cross-Price Elasticity of SSBs for 5^th^ Income Quantile.

| SSBs | Uncompensated Cross Price Elasticity |  |  | Compensated Cross Price Elasticity |  |  |
| --- | --- | --- | --- | --- | --- | --- |
|  | Soft drinks | Tea Outside | Tea Inside | Soft drinks | Tea Outside | Tea Inside |
| Price of Soft drinks (Liter) | - | -0.024<br>(0.041) | -0.163***<br>(0.030) | - | 0.483<br>(0.037) | 0.129***<br>(0.029) |
| Price of Tea Outside (Cup) | 0.081**<br>(0.032) | - | 0.076**<br>(0.030) | 0.398***<br>(0.032) | - | 0.297***<br>(0.029) |
| Price of Tea Inside (KG) | -0.104**<br>(0.042) | 0.173***<br>(0.053) | - | 0.194***<br>(0.043) | 0.521***<br>(0.050) | - |

Notes: \*\*p < 0.05, \*\*\*p < 0.01; Standard errors are in parentheses

## Footnotes

1 Considering 1 USD = 91.7 BDT; Source: World Bank [Official exchange rate BDT/USD downloaded 12/27/2025]

